# Agricultural paraquat dichloride use and Parkinson’s disease in California Central Valley

**DOI:** 10.1101/2022.12.28.22284022

**Authors:** Kimberly C Paul, Myles Cockburn, Yufan Gong, Jeff Bronstein, Beate Ritz

## Abstract

**Background:** Paraquat dichloride is currently among the most widely used commercial herbicides in the United States. Exposure has been broadly linked to Parkinson’s disease (PD) through experimental and epidemiologic research. In the current study, we provide further epidemiologic assessment of ambient paraquat exposure and Parkinson’s risk in a large population-based study of PD in agricultural regions of Central California.

**Methods:** Based on 829 PD patients and 824 community controls, we assessed associations between ambient paraquat dichloride exposure and PD. We estimated residential and workplace proximity to commercial agricultural applications in three California counties since 1974 using the CA pesticide use reporting (PUR) data and land use maps. We evaluated any, duration, and average intensity (pounds per acre per year) of exposure to paraquat in four time-windows prior to PD diagnosis or interview for controls.

**Results:** Ambient paraquat exposure assessed at both residence and workplace was associated with PD based on all three exposure measures, indicating that PD patients lived and worked near agricultural facilities that applied greater amounts of the herbicide than community controls. For workplace proximity to commercial applications since 1974, any exposure (yes/no, OR=1.25, 95% CI=1.00, 1.57), duration of exposure (per SD, OR=1.26, 95% CI=1.10, 1.44), and long-term average intensity of exposure (per SD, OR=1.30, 95% CI=1.10, 1.53) increased the odds of PD. Similar associations were observed for residential proximity (duration of exposure: OR=1.23 per SD, 95% CI=1.07,1.40; long-term average exposure: OR=1.22 per SD, 95% CI=1.03, 1.46). Risk estimates were comparable for men and women and the strongest odds were observed for those diagnosed ≤60 years of age.

**Conclusion:** This study provides further evidence that paraquat dichloride exposure increases the risk of Parkinson’s disease.

## Introduction

Paraquat dichloride, commonly known as paraquat, is currently one of the most widely used commercial herbicides in the United States^1^. It is a quick-acting weed killer also used for desiccation purposes that acts by killing green plant tissue on contact through inhibiting photosynthesis with redox-cycling activity and inducing necrosis^2^. Its great redox-cycling potential has been documented nearly a century, an activity that is also highly toxic to animals and humans^3^. Paraquat (chemical formula [(C6H7N)2]Cl2, aka PQ2+) can undergo cyclic oxidation/reduction reactions, with each cycle generating a highly reactive superoxide radical^4^.

Paraquat exposure was first hypothesized as a possible cause of Parkinson’s disease (PD) in 1987, when an ecologic study in Quebec attributed differences in regional PD prevalence to soil and water contamination from agricultural pesticides, with paraquat being among those most prominently used^5^. Since then, at least ten epidemiologic studies have linked exposure to PD, with a meta-analysis of 13 case-control studies with 3,231 patients and 4,901 controls showing exposure was associated with a 1.64-fold increase in the risk of PD (95% CI=1.27, 2.13)^6^. Epidemiologic results, however, have not been unequivocal and a recent report from the Agricultural Health Study (AHS) suggested no association^7^. Though this contradicts other positive associations reported in the study^8^.

Experimental research has shown paraquat crosses the blood-brain barrier^9,10^ and can enter and accumulate in dopaminergic neurons^11^, the cells lost in PD. Furthermore, studies have demonstrated that paraquat can directly cause and exacerbate α-synuclein pathology^12–16^ and lead to degeneration of dopaminergic neurons^17^, along with other toxic mechanisms. That said, even the first study (Barbeau 1987)^5^ cautioned that it is not necessary for environmental agents to reach the substantia nigra to be toxic to it.

The current study provides a new and expanded epidemiologic assessment of ambient paraquat exposure and PD risk in a large population-based case-control in agricultural Central California, using pesticide use reports and a geographic information system to estimate proximity of residences and workplaces to commercial agricultural applications since 1974.

## Methods

### Study Population

To re-assess paraquat and PD associations, we used both study waves of the Parkinson’s Environment and Genes (PEG) study (n=829 PD patients; n=824 controls). PEG is a population-based case-control study conducted in three agricultural counties in Central California (Kern, Fresno, and Tulare)^18^. Participants were recruited in two waves: wave 1 (PEG1): 2000-2007, n=357 PD patients, n=400 population-based controls; wave 2 (PEG2): 2009-2015, n=472 PD patients, n=424 population-based controls. Patients were enrolled early in disease course (mean PD duration at baseline 3.0 years [SD=2.6]) and all were seen by UCLA movement disorder specialists for in-person neurologic exams, many on multiple occasions, and confirmed as having probable idiopathic PD^19^.

For PEG1 patient enrollment, 1,167 potentially eligible patients were identified through large medical groups, neurologists, and public service announcements, 604 did not meet eligibility criteria for the following reasons: 397 diagnosed with PD >3 years prior to recruitment, 134 lived outside the tri-counties, and 73 did not have PD. Among 563 remaining potential cases, 90 could not be examined by our movement disorder specialists, 56 declined or moved away, and 34 became too ill or died before the scheduled appointment. Of the 473 examined, 94 did not meet criteria for idiopathic PD, an additional 13 were reclassified as not having PD during follow-up, and 6 participants withdrew after examination and before interview. Of the remaining 360 patients, 357 provided all information necessary for inclusion in this study.

For PEG2 patient enrollment, we identified study participants from the pilot PD registry program in California. This registry builds on California laws enacted in 2004 to collect and register PD case records from all health care providers. Between 2010-2015, we screened 2,713 potentially eligible PD patients with an address in the tri-county study area reported to the registry. Of these, 397 denied having PD, 212 were diagnosed with PD prior to 2007 (the earliest diagnosis year allowed for enrollment), 39 did not live in the tri-county area, 1,042 were deceased or too ill, and 293 were unable to be contacted. Of 730 potentially eligible remaining patients, our UCLA movement disorder specialists examined 601 of whom 120 did not meet criteria for idiopathic PD. Of these 481 PD patients, 472 provided all information necessary for inclusion in this study.

Population-based controls for both study waves were required to be living within one of the three counties, have lived in California for at least 5 years before enrollment, not have a diagnosis of PD, and primarily age restricted to ≥55 years of age. Though control age distributions were marginally matched to cases, meaning we did recruit and enroll some controls 35-54. Initially, we identified controls through Medicare enrollee lists (2001), but mostly we selected residents from publicly available residential tax-collector records (after 2001 due to HIPAA restrictions). We used two sampling strategies to increase enrollment success and representativeness of controls for the source population: a) in PEG1, random selection from the Medicare enrollee lists and of residential parcels, followed by mail or phone enrollment; and b) for PEG2, random selection of clustered households (five per cluster, identified from tax-collector records) visited in person by study staff to enroll at least one eligible control from each cluster (only one per household).

For PEG1, we contacted 1,212 potentially eligible controls. Of these individuals, 457 were ineligible: 409 were too young, 44 were too ill to participate, and 4 resided primarily outside the study area. Of 755 eligible population controls, 409 declined participation, were too ill, or moved before an interview was possible, resulting in the enrollment of 346 population controls. A pilot test mailing, for which the number of eligible participants who declined was not known, enrolled another 62 controls. Of these 408 PEG1 controls, 400 provided all information necessary for inclusion in this study. For PEG2, 1,241 potentially eligible controls were contacted, of whom 634 declined participation. Of the 607 PEG2 population controls enrolled, 183 completed only an abbreviated interview that assessed the three most recent residential and workplace addresses, limiting long-term pesticide exposure assessment, thus, these individuals were excluded. From PEG2, 424 controls provided all information necessary for inclusion in this study.

As shown in Table 1, PD patients were on average slightly older than the controls and had a higher proportion of men, European ancestry, and never smokers.

**Table 1.** Study population characteristics and exposure window information.

| **Variable: Mean (SD) or n (%)** | **PD patients (n=829)** | **Controls (n=824)** |
| --- | --- | --- |
| Age | 67.7 (10.6) | 65.9 (11.6) |
| Male Sex | 524 (63.2) | 383 (46.5) |
| Years of Education | 14 (4.0) | 14 (4.0) |
| European Ancestry | 634 (76.5) | 569 (69.2) |
| Non-European Ancestry | 195 (23.5) | 253 (30.8) |
| White | 631 (76.5) | 569 (69.0) |
| Latino | 137 (16.6) | 155 (18.8) |
| Asian | 22 (2.7) | 26 (3.2) |
| Black | 5 (0.6) | 29 (3.5) |
| Other | 30 (3.6) | 45 (5.4) |
| Never Smoker | 449 (54.4) | 397 (48.2) |
| Former Smoker | 345 (41.8) | 331 (40.2) |
| Current Smoker | 31 (3.8) | 96 (11.7) |

### Paraquat Exposure Assessment

We estimated ambient exposure due to living or working near agricultural paraquat dichloride application, using pesticide use record (PUR) based pesticide application data within a geographic information systems (GIS)-based model^20^.

Since 1972, California law mandates the recording of commercial pesticide use in a database maintained by the California Department of Pesticide Regulation (CA-DPR) that includes all commercial agricultural pesticide use by pest control operators and all restricted pesticide use by anyone until 1989, and afterwards (1990-current) all commercial agricultural pesticide use. This database records the location of applications, which can be linked to the Public Land Survey System (PLSS) and the poundage, type of crop, and acreage a pesticide has been applied on, as well as the method and date of application. Figure 1 shows all PUR-reported paraquat application across California and in the tri-county study area since 1974.

**Figure 1.**
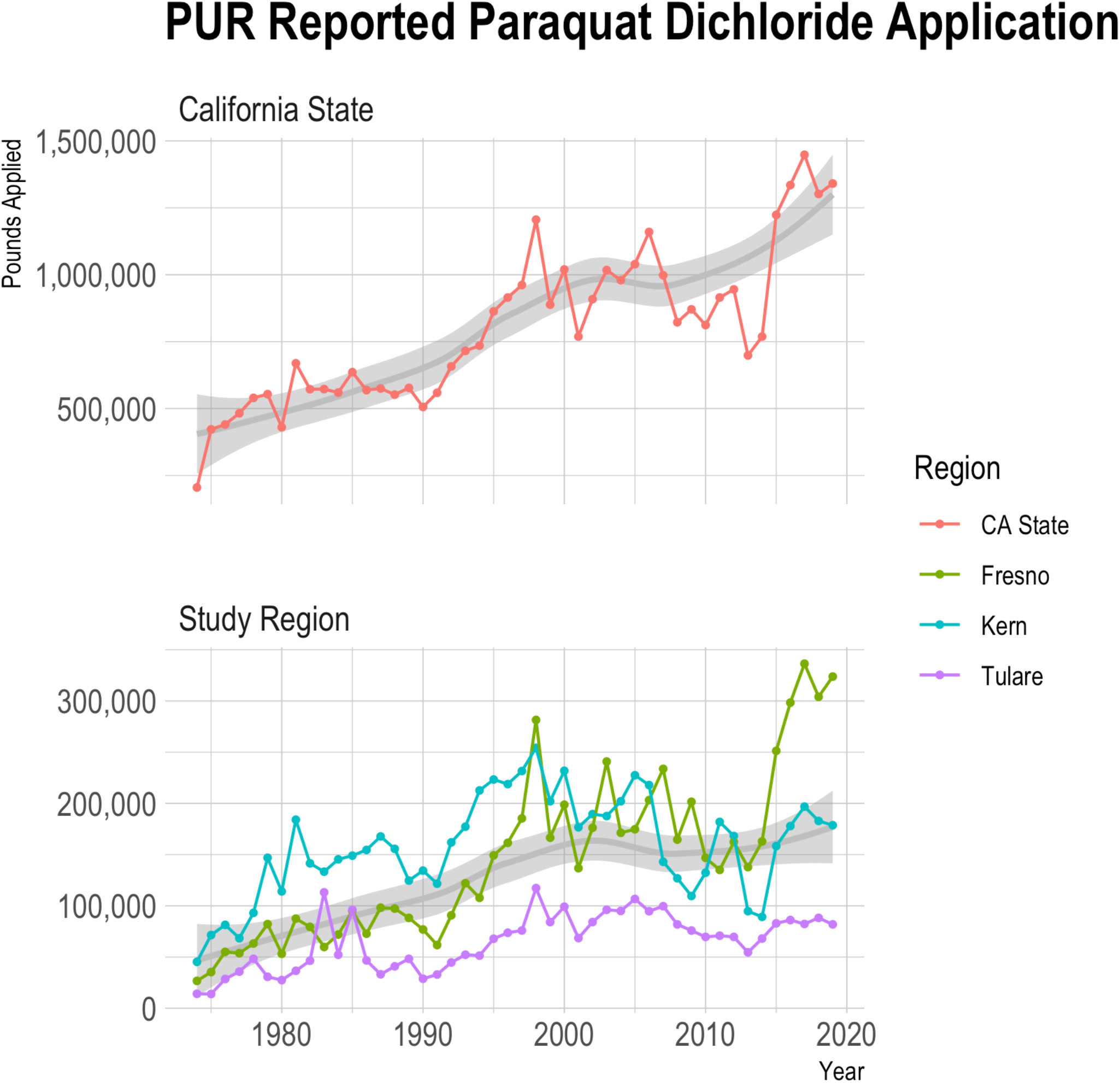
PUR-based paraquat application since 1974 by pounds applied.

We combined the PUR with maps of land-use and crop cover, providing a digital representation of historic land-use, to determine pesticide applications at specific agricultural sites^21^. PEG participants provided lifetime residential and workplace address information, which we geocoded in a multi-step process^22^.

For each pesticide active ingredient, including paraquat dichloride (CA-DPR ChemCode 1601), as well as the commonly used pesticides glyphosate isopropylamine salt (ChemCode 1855), chlorpyrifos (ChemCode 253), and diazinon (ChemCode 198), we determined the pounds of pesticide active ingredient (AI) applied per acre within a 500m buffer of the latitude and longitude representing each residential and workplace address per year since 1974, weighting the total poundage by the proportion of acreage treated (lbs/acre). For our study participants, this resulted in 57,435 annual records for residential and 44,138 occupational records for paraquat exposure with lbs/acre paraquat active ingredient ranging from 0 to 27,211 pounds. We identified several extreme lbs/acre outliers (Supplemental Figure 1) and therefore removed values >99^th^ percentile of the lbs/acre distribution. Figure 2 shows a scatter plot of the data (Figure 2A) and smoothed trend lines based on local regression (Figure 2B).

**Figure 2.**
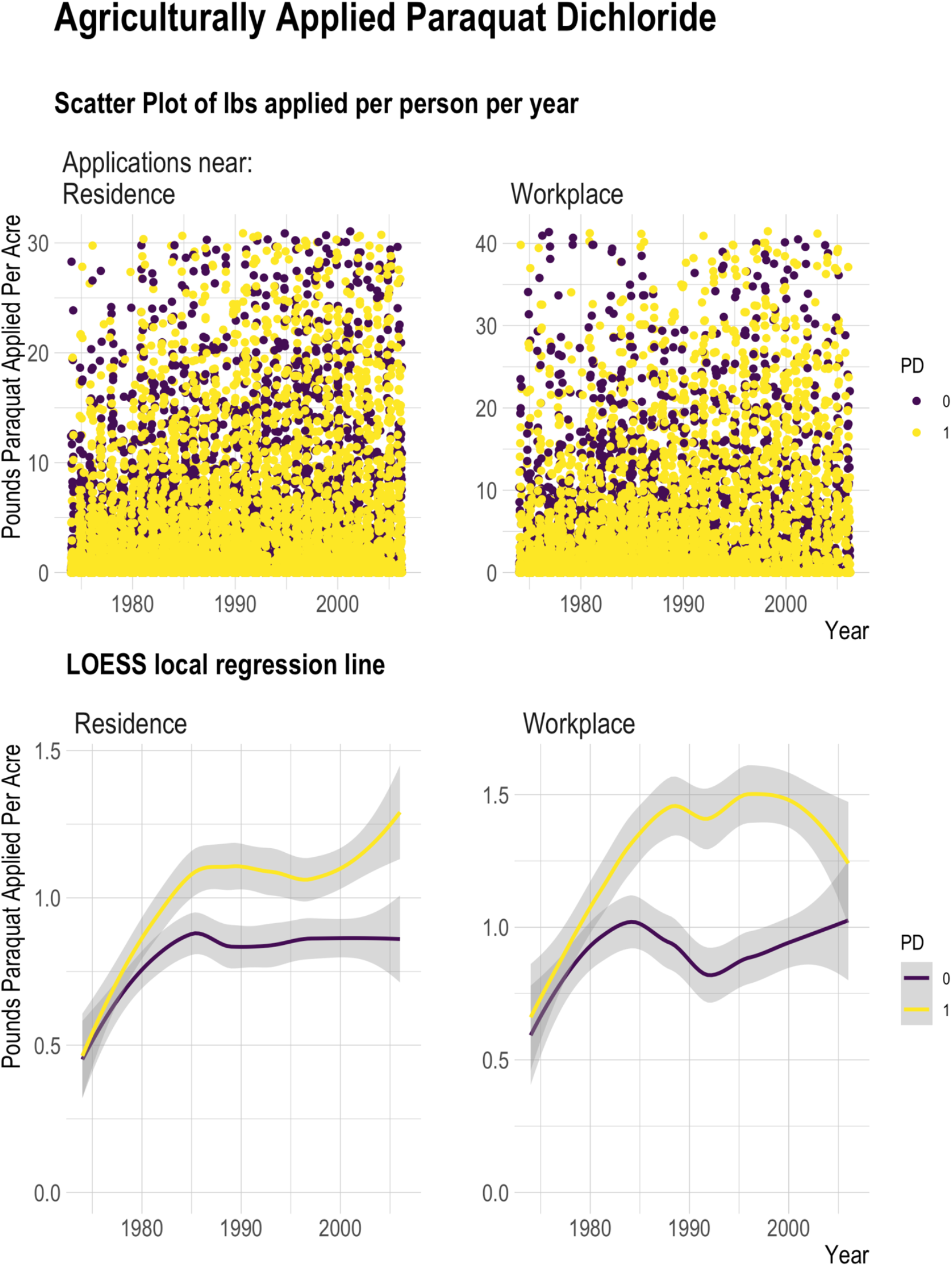
Pounds of agriculturally applied paraquat active ingredient per acre, per subject from 1974-2007. Top: scatter plot of lbs applied/acre for each participant each year within 500m of residential address and workplace address. Bottom: Smoothed trend line from loess local regression based on data shown in plots above.

We report on the following exposure measures for both residential and workplace addresses:

1. Any pesticide application within 500m of the address within the exposure time window (yes/no).
2. Duration of exposure: the number of years participants lived or worked within 500m of agricultural applications as a proportion of the time covered by the window (i.e. # of years with any application in buffer / # of years in window).
3. Average exposure intensity: yearly average pounds of active ingredient applied per acre in the time window (i.e. *lbs Al applied per acre within 500m]/# of years in window*., where AI= active ingredient, y=the first year in the window, and t=last year in the window).

The denominator (# of years in the exposure window) represents the number of years for which the participant provided an address that could be geocoded and linked to the pesticide exposure model (i.e., years with exposure information). We log transformed the average exposure estimates, offset by one, and scaled them to the standard deviations (SD). Duration of exposure is presented as a proportion, scaled to the SD.

We considered four exposure windows for risk assessment: 1) 1974 to index date (PD diagnosis for cases or interview for controls); 2) 1974 to 10 years prior to index date, i.e., lagged 10 years; 3) 20 years to 10 years prior to index date; 4) 10 years prior to index date. By design, the exposure windows covered a very similar length and temporal period on average for patients and controls of each wave (Supplemental Table 1). The mean index year for PD patients was 2006 (SD=4.7 years) and 2006 (SD=3.6 years) for controls. The exposure windows ranged from 8.3 years on average (SD=2.4) to 31.3 years (SD=5.2).

### Statistical Methods

To assess exposure associations, we conducted univariate, unconditional logistic regression to calculate odds ratios (ORs) and 95% confidence intervals (CIs) for PD and estimated paraquat exposure over each time window and each location separately. We controlled for age, gender, race/ethnicity, study wave, and index year (year of diagnosis or interview) to account for temporal trends in pesticide use. We also estimated associations stratified by gender (men and women) and index age (≤60 and > 60 at diagnosis for cases or interview for controls). We conducted several sensitivity analyses to adjust for potential co-exposures. As most participants did not solely live or work near facilities only applying paraquat, we also controlled for the average exposure (same method described above) to chlorpyrifos, glyphosate isopropylamine salt, and diazinon. These three pesticides were selected as they are also widely used agriculturally and/or have been associated with PD in PEG before. We also controlled for occupational use of pesticides and adjusted for the ambient workplace paraquat exposures in the residential models and residential exposures in the workplace models.

## Results

We observed positive associations between paraquat and PD across exposure assessment measures and time-windows (Table 2). For example, for paraquat exposure assessed at residential addresses between 1974 and index year (a 31.3 year exposure window on average), each standard deviation increase in duration of exposure was associated with an 23% increase in the odds of PD (OR=1.23 per SD, 95% CI=1.07, 1.41) and each SD increase average intensity of exposure (average applied pounds per acre) increased the odds by 22% (OR=1.22 per SD, 95% CI=1.03, 1.46). Somewhat stronger associations were observed for workplace proximity to paraquat applications, with higher duration of exposure (OR=1.26 per SD, 95% CI=1.10, 1.44) and average exposure intensity (OR=1.30 per SD, 95% CI=1.10, 1.53) increasing the odds of PD.

**Table 2.** Paraquat risk associations across exposure assessments.

| Exposure Assessment and Time Window | PD patients<br>(n=824) | Controls<br>(n=820) | OR (95% CI) | p-value | PD patients<br>(n=800) | Controls<br>(n=781) | OR (95% CI) | p-value |
| --- | --- | --- | --- | --- | --- | --- | --- | --- |
|  | n (%) or Mean (SD) |  |  |  | n (%) or Mean (SD) |  |  |  |
| Application Near: | Residence |  |  |  | Workplace |  |  |  |
| Any application, yes |  |  |  |  |  |  |  |  |
| 1974-index year | 591 (71.7) | 576 (70.2) | 1.12 (0.90, 1.41) | 0.31 | 551 (68.9) | 499 (63.9) | 1.25 (1.00, 1.57) | 0.05 |
| 1974-index year with 10y lag | 561 (68.2) | 537 (65.6) | 1.16 (0.93, 1.45) | 0.18 | 536 (67.1) | 467 (59.8) | 1.33 (1.07, 1.66) | 0.01 |
| 20y - 10y prior to index year | 423 (51.6) | 363 (44.4) | 1.34 (1.08, 1.65) | 0.007 | 382 (49.9) | 305 (42.8) | 1.33 (1.06, 1.67) | 0.01 |
| 10y prior to index year | 290 (35.5) | 347 (42.3) | 1.33 (1.07, 1.64) | 0.01 | 201 (33.9) | 230 (34.2) | 1.10 (0.85, 1.42) | 0.48 |
| Duration of exposure, per SD |  |  |  |  |  |  |  |  |
| 1974-index year | 0.23 (0.28) | 0.19 (0.24) | 1.23 (1.07, 1.41) | 0.003 | 0.24 (0.29) | 0.18 (0.24) | 1.26 (1.10, 1.44) | 0.001 |
| 1974-index year with 10y lag | 0.25 (0.30) | 0.21 (0.26) | 1.22 (1.07, 1.40) | 0.004 | 0.26 (0.31) | 0.20 (0.26) | 1.27 (1.10, 1.46) | 0.0008 |
| 20y - 10y prior to index year | 0.25 (0.33) | 0.19 (0.30) | 1.23 (1.08, 1.40) | 0.002 | 0.25 (0.34) | 0.18 (0.29) | 1.30 (1.14, 1.50) | 1.62E-04 |
| 10y prior to index year | 0.20 (0.31) | 0.16 (0.28) | 1.17 (1.04, 1.33) | 0.01 | 0.19 (0.33) | 0.15 (0.27) | 1.21 (1.05, 1.39) | 0.008 |
| Average exposure, per SD |  |  |  |  |  |  |  |  |
| 1974-index year | 0.46 (0.62) | 0.39 (0.56) | 1.22 (1.03, 1.46) | 0.02 | 0.49 (0.69) | 0.37 (0.60) | 1.30 (1.10, 1.53) | 0.002 |
| 1974-index year with 10y lag | 0.44 (0.61) | 0.38 (0.56) | 1.24 (1.04, 1.48) | 0.02 | 0.48 (0.67) | 0.36 (0.60) | 1.28 (1.08, 1.52) | 0.005 |
| 20y - 10y prior to index year | 0.44 (0.71) | 0.35 (0.62) | 1.23 (1.06, 1.45) | 0.008 | 0.48 (0.78) | 0.32 (0.64) | 1.35 (1.15, 1.58) | 2.21E-04 |
| 10y prior to index year | 0.40 (0.72) | 0.32 (0.65) | 1.17 (1.00, 1.37) | 0.04 | 0.42 (0.83) | 0.30 (0.69) | 1.25 (1.07, 1.47) | 0.006 |

Overall, most of the study population in this agricultural region both lived and worked within 500m of at least one paraquat application since 1974, though the proportion was higher among patients than controls (residence: 72% of patients and 70% of controls; workplace: 69% of patients and 64% of controls; Table 2). However, a much higher proportion of patients lived near facilities applying paraquat in the 10 years prior to index than controls (42% versus 36%; OR=1.33, 95% CI=1.07, 1.64) and between 20 and 10 years prior to index (52% versus 44%; OR=1.34, 95% CI=1.08, 1.65). A higher proportion of patients also worked near paraquat applications than controls in all windows, including the period 20 to 10 years prior to diagnosis (50% versus 43%; OR=1.33, 95% CI=1.06, 1.67). In this time window, patients lived near agricultural fields or facilities applying paraquat for 25% of the years on average compared with the controls’ 19% (OR=1.23 per SD, 95% CI=1.08, 1.40). Furthermore, the patients also worked near commercial paraquat applications for 25% of the years in this time-period on average compared with the controls’ 18% on average (OR=1.30 per SD, 95% CI=1.14, 1.50; Table 2). Similar results were observed for other time windows and for average exposure intensity (pounds applied per acres).

Risk estimates stratified by gender were generally similar, with somewhat stronger associations from workplace exposures in men (Table 3). For example, at workplace addresses, for duration we estimated an odds ratio of 1.31 among men (95% CI=1.10, 1.57) and 1.20 (95% CI=0.96, 1.50) for women. For age at index date, risk estimates were higher among those ≤60 than >60 years of age. Increased duration of exposure at residential addresses from 1974 to index year increased the odds 1.64-fold per SD among those ≤60 (95% CI=1.18, 2.28) and 1.16-fold among those >60 years (95% CI=1.01, 1.36). Such differences were also observed for other exposure measures and time windows.

**Table 3.**
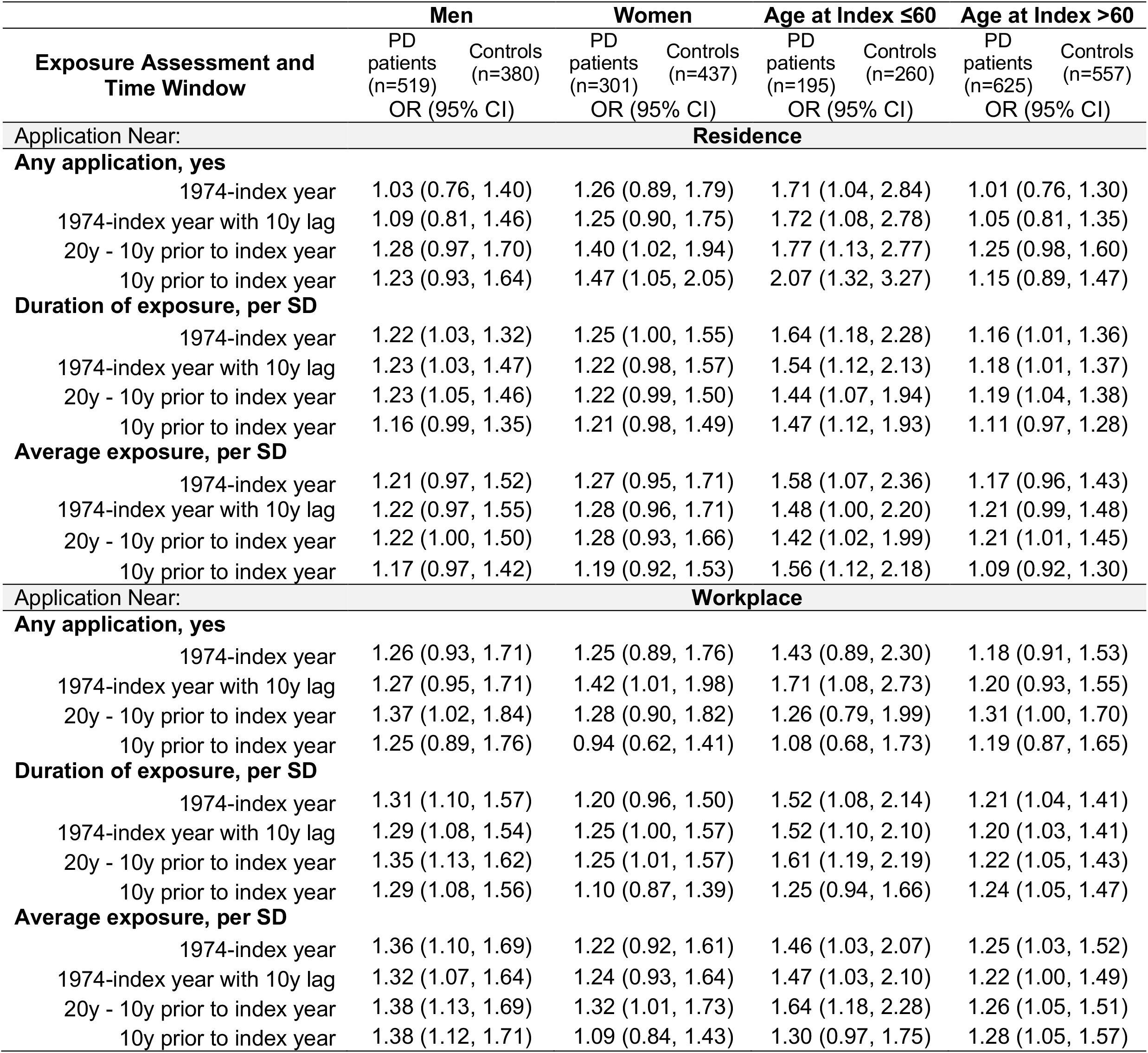
Paraquat risk associations across exposure assessments: stratified estimates.

The odds ratios were generally robust to including measures of other pesticide exposures in the models, including adding an indicator for occupational pesticide use, adjusting for residential or workplace proximity to chlorpyrifos, diazinon, and glyphosate applications, and adjusting for ambient workplace paraquat exposures in the residential models and residential exposures in the workplace models (Table 4). Though there was some attenuation of residential paraquat odds ratios when including the ambient workplace exposures and for some but not all residential exposure estimates when adjusting for proximity to chlorpyrifos, diazinon, and glyphosate applications.

**Table 4.** Paraquat risk associations across exposure assessments. Multi-pesticide adjusted models.

| Exposure Assessment and Time Window | Adjusted for any reported occupational pesticide use |  | Adjusted for ambient chlorpyrifos, diazinon, and glyphosate |  | Adjusted for paraquat at other location |  |
| --- | --- | --- | --- | --- | --- | --- |
|  | Residence | Workplace | Residence | Workplace | Residence | Workplace |
|  | OR (95% CI) | OR (95% CI) | OR (95% CI) | OR (95% CI) | OR (95% CI) | OR (95% CI) |
| **Any application, yes** |  |  |  |  |  |  |
| 1974-index year | 1.10 (0.87, 1.38) | 1.21 (0.97, 1.52) | 0.99 (0.77, 1.27) | 1.14 (0.89, 1.46) | 1.04 (0.81, 1.34) | 1.22 (0.96, 1.56) |
| 1974-index year with 10y lag | 1.13 (0.91, 1.41) | 1.29 (1.04, 1.62) | 1.03 (0.81, 1.32) | 1.23 (0.97, 1.57) | 1.05 (0.82, 1.65) | 1.29 (1.01, 1.65) |
| 20y - 10y prior to index year | 1.30 (1.05, 1.61) | 1.29 (1.03, 1.63) | 1.16 (0.90, 1.49) | 1.22 (0.93, 1.60) | 1.05 (0.82, 1.34) | 1.29 (1.01, 1.65) |
| 10y prior to index year | 1.28 (1.03, 1.60) | 1.07 (0.82, 1.38) | 1.14 (0.88, 1.48) | 0.96 (0.72, 1.30) | 1.44 (1.11, 1.89) | 0.95 (0.72, 1.26) |
| **Duration of exposure, per SD** |  |  |  |  |  |  |
| 1974-index year | 1.19 (1.04, 1.37) | 1.23 (1.07, 1.41) | 1.14 (0.94, 1.39) | 1.23 (1.01, 1.49) | 1.10 (0.93, 1.31) | 1.18 (0.99, 1.40) |
| 1974-index year with 10y lag | 1.19 (1.04, 1.37) | 1.24 (1.08, 1.42) | 1.14 (0.95, 1.37) | 1.24 (1.02, 1.50) | 1.09 (0.92, 1.42) | 1.19 (1.00, 1.42) |
| 20y - 10y prior to index year | 1.20 (1.05, 1.36) | 1.27 (1.11, 1.46) | 1.13 (0.95, 1.35) | 1.35 (1.12, 1.64) | 1.09 (0.92, 1.29) | 1.19 (1.00, 1.42) |
| 10y prior to index year | 1.14 (1.01, 1.30) | 1.18 (1.03, 1.37) | 1.06 (0.91, 1.25) | 1.20 (1.01, 1.43) | 1.13 (0.97, 1.32) | 1.15 (0.99, 1.34) |
| **Average exposure, per SD** |  |  |  |  |  |  |
| 1974-index year | 1.18 (0.99, 1.41) | 1.27 (1.07, 1.50) | 1.07 (0.84, 1.36) | 1.20 (0.95, 1.51) | 1.05 (0.91, 1.38) | 1.19 (1.03, 1.38) |
| 1974-index year with 10y lag | 1.20 (1.00, 1.43) | 1.25 (1.05, 1.49) | 1.03 (0.80, 1.32) | 1.23 (0.98, 1.55) | 1.06 (0.92, 1.23) | 1.17 (1.01, 1.36) |
| 20y - 10y prior to index year | 1.21 (1.03, 1.41) | 1.32 (1.12, 1.55) | 1.08 (0.88, 1.33) | 1.39 (1.13, 1.73) | 1.06 (0.92, 1.36) | 1.17 (1.01, 1.36) |
| 10y prior to index year | 1.14 (0.98, 1.33) | 1.22 (1.04, 1.45) | 1.02 (0.84, 1.23) | 1.24 (1.03, 1.50) | 1.07 (0.93, 1.23) | 1.18 (1.03, 1.37) |

## Discussion

In this population-based case-control study of Parkinson’s disease, we investigated ambient paraquat exposure estimated via residential and workplace proximity to agricultural paraquat application. Paraquat was associated with an increased risk of PD with multiple exposure assessment measures (any exposure, duration of exposure, and average exposure intensity) and across multiple exposure windows, ranging from 8 to 31 years on average. Overall, a higher proportion of the PD patients both lived and worked near commercial paraquat application, which were primarily related to agriculture in our study region, than controls. Applications were on average greater in terms of the amount applied (pounds of active ingredient applied per acre) and took place over a longer duration (proportion of years in the time windows with exposure). Exposure associations were strongest for younger-onset patients (≤60 at diagnosis)^23^, replicating previous results seen in the PEG1 study with data from the PEG2 study wave. Furthermore, while risks were increased in all exposure windows, exposures decades prior to diagnosis, such as twenty and thirty years prior, were associated with the strongest risks.

This research adds to an existing body of literature that previously connected the herbicide to PD. Paraquat was initially scrutinized for its potential to cause PD due to its structural similarity to MPP+, the toxic metabolite of MPTP, which was found to induce parkinsonism in humans in 1983^24^. Since then, at least 13 case-control studies and one prospective cohort have investigated paraquat exposure. A recent systematic review and meta-analysis reported a summary estimate for PD and paraquat in 13 case control studies of OR = 1.64 (95% CI=1.27, 2.13)^6^. At the same time, experimental research has linked exposure to dopaminergic neuron toxicity and α-synuclein related biology among other mechanisms^9–17^. Still, epidemiologic results, have not been unanimous and a recent report from the longitudinal Agricultural Health Study (AHS) has suggested no association^7^. Loss to follow-up however may have affected the AHS results, as identification of PD cases mainly depended on self-report and only 46% of initially enrolled male farmers had not dropped out by the end of follow-up^7^. The AHS cohort study results also contradict strongly positive associations reported for paraquat and PD by the FAME-PD case-control study nested within the AHS^8^, leaving open questions about accurate self-reporting of pesticide use and selection bias in these studies.

Our study makes use of agricultural pesticide application records required by law in California to determine the amount of paraquat active ingredient applied within 500m of participants lifetime reported residential and workplace addresses. This allowed us to assess exposures to applications as far back as 1974 without relying on participant recall. This is especially important for PD, where distant exposures decades prior to diagnosis are likely relevant. With our record-based exposure assessment, we were also able to greatly minimize recall bias and exposure misclassification, while still investigating historic exposures. Furthermore, PD is a commonly misdiagnosed disorders, with estimates of misdiagnosis often ranging from 20-30%^25^. All of our patients were seen in person by the same UCLA movement disorder specialists to confirm diagnosis through clinical characteristics. Thus, also minimizing outcome misclassification.

Historically, paraquat has been applied by ground, aerial, and backpack methods. It is highly persistent in the soil environment, with field studies reporting a half-life ranging from 1.4 to 7.2 years^26^. It also strongly binds to certain soils, which limits the threat of groundwater contamination^26^. But this property may also increase the risk of contaminated soil being blown or tracked to nearby locations, such as homes. Furthermore, prior to US EPA restrictions of aerial applications and during the exposure windows of interest for the PEG study, paraquat has been detected in airborne samples up to 1600m from single field aerial applications in the San Joaquin Valley in California^27,28^. Other studies of paraquat drift have demonstrated foliage damage extending over a ½ mile from fields into a neighboring community^29^. Our exposure buffer was limited to 500m. We previously conducted a validation study demonstrating that PUR model-derived organochlorine estimates of exposure within 500m of homes had high specificity in predicting blood-measured DDE levels^30^. Though it should be noted, our exposure assessment method did not account for potentially relevant factors, such as wind patterns at the time of application or geographic features that may influence pesticide drift. However, long term exposures from contaminated dusts may depend less on the wind direction on the application day and the geography of the central valley areas, with the most intensive agriculture regions homogeneously flat. Our method also assumes that the participant was at the recorded location during the exposure relevant time or that the agent was still active and exposed the residents after application had occurred. Exposure misclassification, however, would likely be non-differential to case status and therefore bias results toward the null.

Additionally, the reality of commercial agriculture is that many different pesticides may be applied on the same fields in seasonal patterns year after year. Thus, participants with residential or workplace proximity to paraquat application also lived and worked in proximity to other pesticide applications. We performed a series of co-adjustments for other commonly applied pesticides, including chlorpyrifos, diazinon, and glyphosate. While there was some attenuation of risk estimates, particularly for some of the residential exposures, paraquat was consistently positively associated with Parkinson’s disease for most of our exposure measures and windows. In fact, duration and average intensity of exposure at workplace even showed stronger associations in multi-pesticide adjusted models. As real-world co-exposures are the norm and not the exception due to intensive and changing agricultural pesticide use, co-exposure mixtures should be investigated to evaluate their toxicity as a part of policy and regulation.

Overall, this study provides further evidence that paraquat exposure increases the risk of Parkinson’s disease.

## Data Availability

Data produced in the present study are available upon reasonable request to the authors

## References

1. US EPA. Paraquat Dichloride. https://www.epa.gov/ingredients-used-pesticide-products/paraquat-dichloride 2022.

2. Bus JS, Gibson JE. Paraquat: model for oxidant-initiated toxicity. Environ Health Perspect. 1984;55.

3. Dinis-Oliveira RJ, Duarte JA, Sánchez-Navarro A, Remião F, Bastos ML, Carvalho F. Paraquat poisonings: Mechanisms of lung toxicity, clinical features, and treatment. Crit Rev Toxicol 2008.

4. Peterson ME, Talcott PA. Small Animal Toxicology, Third Edition. Small Animal Toxicology, Third Edition 2012.

5. Barbeau A, Roy M, Bernier G, Campanella G, Paris S. Ecogenetics of Parkinson’s Disease: Prevalence and Environmental Aspects in Rural Areas. Canadian Journal of Neurological Sciences / Journal Canadien des Sciences Neurologiques. 1987;14(1).

6. Tangamornsuksan W, Lohitnavy O, Sruamsiri R, et al. Paraquat exposure and Parkinson’s disease: A systematic review and meta-analysis. Arch Environ Occup Health. 2019;2019(5).

7. Shrestha S, Parks CG, Umbach DM, et al. Pesticide use and incident Parkinson’s disease in a cohort of farmers and their spouses. Environ Res. 2020;191.

8. Tanner CM, Kame F, Ross GW, et al. Rotenone, paraquat, and Parkinson’s disease. Environ Health Perspect. 2011;

9. McCormack AL, Monte DA di. Effects of L-dopa and other amino acids against paraquat-induced nigrostriatal degeneration. J Neurochem. 2003;2003(1).

10. Shimizu K, Ohtaki K, Matsubara K, et al. Carrier-mediated processes in blood-brain barrier penetration and neural uptake of paraquat. Brain Res. 2001;906(1–2).

11. Rappold PM, Cui M, Chesser AS, et al. Paraquat neurotoxicity is mediated by the dopamine transporter and organic cation transporter-3. Proc Natl Acad Sci U S A. 2011;2011(51).

12. Uversky VN, Li J, Fink AL. Pesticides directly accelerate the rate of α-synuclein fibril formation: A possible factor in Parkinson’s disease. FEBS Lett. 2001;2001(3).

13. Manning-Bog AB, McCormack AL, Li J, Uversky VN, Fink AL, Monte DA di. The herbicide paraquat causes up-regulation and aggregation of α-synuclein in mice: Paraquat and α-synuclein. Journal of Biological Chemistry. 2002;2002(3).

14. Wills J, Credle J, Oaks AW, et al. Paraquat, but not maneb, induces synucleinopathy and tauopathy in striata of mice through inhibition of proteasomal and autophagic pathways. PLoS One. 2012;2012(1).

15. Naudet N, Antier E, Gaillard D, et al. Oral exposure to paraquat triggers earlier expression of phosphorylated α-synuclein in the enteric nervous system of A53T mutant human α-synuclein transgenic mice. J Neuropathol Exp Neurol. 2017;2017(12).

16. Musgrove RE, Helwig M, Bae EJ, et al. Oxidative stress in vagal neurons promotes parkinsonian pathology and intercellular α-synuclein transfer. Journal of Clinical Investigation. 2019;2019(9).

17. McCormack AL, Thiruchelvam M, Manning-Bog AB, et al. Environmental risk factors and Parkinson’s disease: Selective degeneration of nigral dopaminergic neurons caused by the herbicide paraquat. Neurobiol Dis. 2002;2002(2).

18. Ritz BR, Paul KC, Bronstein JM. Of Pesticides and Men: a California Story of Genes and Environment in Parkinson’s Disease. Curr Environ Health Rep. 2016;2016(1).

19. Hughes AJ, Ben-Shlomo Y, Daniel SE, Lees a J. What features improve the accuracy of clinical diagnosis in Parkinson’s disease: a clinicopathologic study. Neurology. 1992;42(6):1142–1146.

20. Cockburn M, Mills P, Zhang X, Zadnick J, Goldberg D, Ritz B. Prostate cancer and ambient pesticide exposure in agriculturally intensive areas in California. Am J Epidemiol. 2011;173(11):1280–1288.

21. CDWR. California Department of Water Resources Land Use Surveys [Internet]. 2013 [cited 2015 Jul 27]. Available from: http://www.water.ca.gov/landwateruse/lusrvymain.cfm

22. McElroy JA, Remington PL, Trentham-Dietz A, Robert SA, Newcomb PA. Geocoding addresses from a large population-based study: Lessons learned. Epidemiology. 2003;14(4):399–407.

23. Costello S, Cockburn M, Bronstein J, Zhang X, Ritz B. Parkinson’s disease and residential exposure to maneb and paraquat from agricultural applications in the central valley of California. Am J Epidemiol. 2009;169(8):919–926.

24. William Langston J, Ballard P, Tetrud JW, Irwin I. Chronic parkinsonism in humans due to a product of meperidine-analog synthesis. Science (1979). 1983;219(4587).

25. Poewe W, Wenning G. The differential diagnosis of Parkinson’s disease. Eur J Neurol 2002.

26. Roede JR, Miller GW. Paraquat. Encyclopedia of Toxicology: Third Edition. 2014.

27. Chester G, Ward RJ. Occupational exposure and drift hazard during aerial application of paraquat to cotton. Arch Environ Contam Toxicol. 1984;1984(5).

28. Seiber JN, Woodrow JE. Sampling and analysis of airborne residues of paraquat in treated cotton field environments. Arch Environ Contam Toxicol. 1981;1981(2).

29. Ames RC. Community exposure to a paraquat drift. Arch Environ Health. 1993;1993(1).

30. Ritz B, Costello S. Geographic model and biomarker-derived measures of pesticide exposure and Parkinson’s disease. Ann N Y Acad Sci. 2006. p. 378–387.

