## Supplemental Materials for "Agricultural paraquat dichloride use and Parkinson’s disease in California Central Valley"

**Supplemental Table 1. Exposure window information in PEG study (n=1653)**

| | PD patients (n=829) | | Controls (n=824) | |
|---|---|---|---|---|
| | n | Mean (SD) | n | Mean (SD) |
| **Exposure Window Information** | | | | |
| Index year | 829 | 2006 (4.7) | 824 | 2006 (3.6) |
| 10 years prior to Index year | 829 | 1996 (4.7) | 824 | 1996 (3.6) |
| 20 years prior to Index year | 829 | 1986 (4.7) | 824 | 1986 (3.6) |
| **Average Years of Exposure Address History** | | | | |
| Residence | | | | |
| 1974-index year | 824 | 31.3 (5.2) | 820 | 31.3 (4.0) |
| 1974-index year with 10-year lag | 823 | 21.6 (4.9) | 819 | 21.6 (3.8) |
| 20 years - 10 years prior to index year | 820 | 9.8 (0.8) | 818 | 9.9 (0.8) |
| 10 years prior to index year | 820 | 9.8 (0.9) | 817 | 9.8 (0.8) |
| Workplace | | | | |
| 1974-index year | 800 | 27.1 (9.0) | 781 | 23.8 (9.1) |
| 1974-index year with 10-year lag | 799 | 19.7 (6.4) | 781 | 17.5 (6.5) |
| 20 years - 10 years prior to index year | 765 | 9.2 (1.2) | 712 | 8.7 (2.3) |
| 10 years prior to index year | 672 | 8.8 (2.4) | 593 | 8.3 (2.7) |

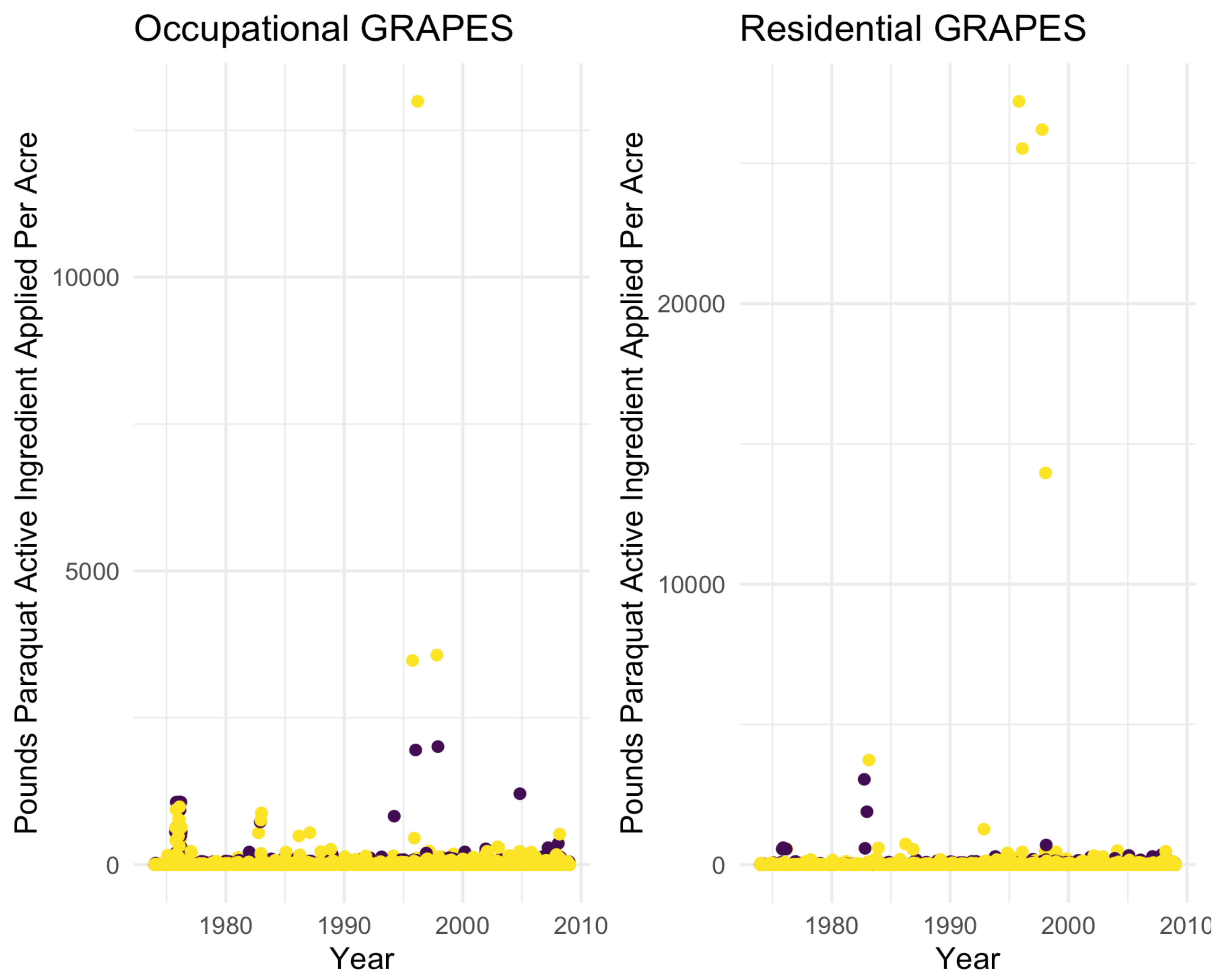


**Supplemental Figure 1.** Plot of all lbs/acre estimates for paraquat across all years and participants. Yellow=patients; purple=controls
